# Distinct Associations of Gait Variability and Physical Fitness with Resting-State Dynamic Functional Connectivity in Cognitively Normal Older Adults from the AGUEDA Trial

**DOI:** 10.64898/2026.11.25.26368939

**Authors:** Andrea Coca-Pulido, Patricio Solis-Urra, Isa Martín-Fuentes, Oren Contreras-Rodríguez, Emilio José Ruiz-Malagón, Elena de la Calle, Michelle W. Voss, Jose Mora-Gonzalez, Irene Esteban-Cornejo

## Abstract

Gait variability and physical fitness may provide accessible indicators of brain health in aging, yet their associations with resting-state dynamic functional connectivity remain unclear. We examined associations of gait variability and physical fitness with local and distant temporal consistency of resting-state dynamic functional connectivity in 86 cognitively normal older adults from the AGUEDA trial. Gait variability was assessed using coefficients of variation for stride length, gait velocity, step time, and step length, while physical fitness included muscular strength and cardiorespiratory fitness. Dynamic functional connectivity was estimated using voxel-wise stepwise connectivity analyses. Analyses targeting regions related to Alzheimer’s disease risk and exploratory whole-brain analyses were adjusted for age, sex, and education. Greater gait variability was associated with local, but not distant, temporal consistency of dynamic functional connectivity. Greater stride length, gait velocity, and step length variability were associated with higher local temporal consistency in the left inferior frontal gyrus (β = 0.28–0.33; false discovery rate-corrected P = 0.024–0.047) and lower temporal consistency in the right insula (β = −0.47 to −0.31; false discovery rate-corrected P = 0.001–0.028). Whole-brain analyses revealed lower local temporal consistency in the left parahippocampal gyrus with greater stride length variability (β = −0.43; family-wise error-corrected P = 0.002) and in the left calcarine cortex with greater gait velocity variability (β = −0.45; family-wise error-corrected P = 0.001). No statistically significant associations were observed for physical fitness. These findings suggest that gait variability may reflect aspects of local dynamic functional brain organization in cognitively normal older adults.

**Abbreviated Summary:** Coca-Pulido et al. investigated associations of gait variability and physical fitness with dynamic functional brain connectivity in cognitively normal older adults. Greater gait variability was associated with region-specific patterns of local connectivity, whereas no significant associations were found for physical fitness or distant connectivity.

## 1. Introduction

Aging is accompanied by marked interindividual variability in physical performance indicators, including gait variability, muscular strength, and cardiorespiratory fitness (CRF) ^1^. These dimensions of physical performance are closely related to cognitive processes and rely on overlapping neural substrates, particularly within prefrontal, parietal, sensorimotor and cerebellar regions that support motor control and executive functioning ^2–4^. Consequently, age-related declines in physical performance indicators—such as increased gait variability or reduced muscular strength and CRF—have been increasingly recognized as sensitive markers of underlying brain health in older adults ^5,6^.

Most neuroimaging studies examining gait variability and physical fitness indicators have focused on resting-state functional connectivity (rs-FC) using static approaches. Poorer stability as measured by greater gait variability is consistently associated with weaker connectivity within and between frontoparietal, sensorimotor, and attention networks, suggesting reduced efficiency of motor–cognitive integration ^2,4,7^. By comparison, higher muscular strength and CRF have been linked to better static rs-FC across large-scale networks, including the default mode, frontoparietal, and sensorimotor systems, which support both cognitive and motor performance ^8–11^. Together, these findings indicate that physical performance indicators are closely related to the integrity of intrinsic brain networks in aging.

Static rs-FC captures the average strength of connectivity over time and does not reflect the dynamic nature of brain communication. Neural activity is inherently dynamic, with functional connections continuously reconfiguring to support adaptive behavior. Resting-state dynamic functional connectivity (rs-dFC) provides a complementary perspective by characterizing how consistently brain regions remain functionally coupled across time (i.e., the recurrence of coupling across time rather than an average estimate across the entire scan, which may reflect either sustained coupling or transient periods of stronger coupling)^12–14^. This approach also allows the distinction between local and distant functional interactions, thereby capturing the temporal consistency of both short-range and long-range coordination across the brain at rest. Importantly, reduced rs-dFC temporal consistency has been associated with poorer cognitive and motor performance in older adults ^15^. In addition, recent evidence from our group showed that rs-dFC patterns in cognitively normal older adults are related to multiple AD risk markers, supporting the sensitivity of this approach to early alterations linked to AD vulnerability even before cognitive symptoms emerge ^16^. Together, these findings suggest that rs-dFC may provide a particularly sensitive marker of age-related differences in the temporal organization of regional coordination at rest. Despite growing evidence linking gait variability and physical fitness indicators to static rs-FC, no previous studies have examined their associations with rs-dFC. Understanding how physical performance indicators relate to the temporal consistency of local and distant functional interactions may therefore provide novel insights into the neural mechanisms supporting motor and cognitive function in later life, beyond what static measures can reveal. Moreover, examining these associations within regions previously linked to AD risk markers may help determine whether physical performance indicators capture variation in functional brain organization that is relevant to early AD-related vulnerability, even in cognitively normal aging.

Therefore, the present study aimed to examine cross-sectional associations between gait variability and physical fitness indicators (muscular strength and CRF) and rs-dFC temporal consistency in cognitively normal older adults. Given prior evidence that rs-dFC is sensitive to early AD-related alterations even in cognitively normal populations, two complementary approaches were applied: (1) a targeted analysis focused on AD risk-related brain regions, and (2) a whole-brain exploratory analysis. Based on previous studies linking poorer gait and physical performance to altered functional connectivity in motor-cognitive systems, we hypothesized that greater gait variability and poorer physical fitness indicators would be associated with lower local rs-dFC temporal consistency in regions supporting motor and executive control, reflecting less stable short-range functional coordination during aging.

## 2. Material and Methods

### 2.1. Study design and participants

A total of 91 cognitively normal, physically inactive older adults aged 65–80 years were recruited to participate in the “Active Gains in brain Using Exercise During Aging” (AGUEDA) trial (ClinicalTrials.gov: (NCT05186090)), a randomized controlled trial evaluating a 24-week resistance exercise program on executive function ^17^. Participants were recruited from Granada (Spain). Full inclusion and exclusion criteria have been reported elsewhere ^18^. Briefly, participants were required to be physically inactive and cognitively healthy, with no relevant depressive symptoms. All instruments were administered in validated Spanish versions. For this cross-sectional analysis, we used baseline data collected between March 2021 and May 2022. As this study involved a secondary cross-sectional analysis of baseline data from the AGUEDA trial, no study-specific sample size calculation was performed. The analytical sample was determined by the availability of valid baseline gait variability, physical fitness, and neuroimaging data.

After quality control, five participants were excluded due to excessive head motion or invalid resting-state fMRI slices. In addition, four participants were excluded from gait-related analyses due to unavailable or invalid gait variability data. One participant had missing data for neurofilament light (NfL) and two participants had missing data for APOE4 genotyping. As a result, gait-related analyses were conducted with up to 81 participants, and physical fitness analyses with up to 86 participants. The study was conducted in accordance with the Declaration of Helsinki and approved by the Ethics Board of the Andalusian Health Service (CEIM/CEI Provincial de Granada; #2317-N-19). Written informed consent was obtained from all participants.

### 2.2. Gait variability indicators

Gait variability was assessed using the OptoGait system (Microgate Srl; Bolzano, Italy). Participants walked at their usual pace across a 5.4-m walkway equipped with transmitting and receiving bars containing photoelectric sensors that detect footfall timing and position. From 30 valid footsteps per participant, the coefficient of variation (CV) for stride length, gait velocity, step time, and step length was calculated to capture variability across consecutive steps. Lower values of gait variability indicators indicate better performance and better gait stability. Further methodological details are available elsewhere ^19^.

### 2.3. Physical performance indicators

Physical performance was assessed through indicators of muscular strength and CRF. Muscular strength was evaluated using handgrip strength, biceps curl strength, squats strength, and isokinetic strength (elbow and knee extension strength). CRF was assessed using the 2-km walking test and the 6-min walking test. For all physical fitness indicators, higher values reflect better performance, except for the 2-km walking test, where shorter completion times indicate better performance; this variable was inverted for the analyses so that higher values consistently represented better function. More detailed information about the assessment procedures and specific tests is available elsewhere ^20^.

### 2.4. Resting-State Dynamic Functional Connectivity (rs-dFC)

Resting state-fMRI data were acquired on a 3.0 T Siemens Magnetom Prisma Fit scanner (Siemens Medical Solutions, Erlangen, Germany) with a 64-channel head coil (multiband EPI sequence, TR = 1000 ms, TE = 40 ms, voxel size = 2.5 mm³, scan duration = 8:12 min), together with high-resolution T1-weighted images (TR = 2400 ms, TE = 2.31 ms, inversion time = 1060 ms, field of view = 256 mm, voxel size = 0.8 × 0.8 × 0.8 mm, 224 slices, and scan duration = 6 min and 38 s). Preprocessing was performed in Neurodesk^21^ using FSL, Matlab, and CONN. Distortion correction with fieldmaps (topup), realignment, MNI normalization, spatial smoothing (6 mm FWHM), and band-pass filtering (0.008–0.09 Hz) were applied. Subjects with more than 25% invalid resting-state fMRI volumes, identified during the default CONN quality-control/scrubbing procedure, were excluded from the analyses. Average framewise displacement was examined descriptively and was elevated in all excluded participants (>0.60 mm).

Rs-dFC reflects how consistently functional interactions between brain regions are maintained over time, based on correlations in their resting-state time series, thereby providing a measure of the temporal consistency of regional coordination during rest. Connectivity patterns were characterized as local (short-range) or distant (long-range), reflecting the spatial extent of coupling within the brain’s topological organization. In the context of network neuroscience, local FC reflects coupling between nodes that are strongly and directly functionally related, often within the same network, whereas distant FC reflects coupling between nodes linked through indirect paths, often across different networks. To compute these maps, a sliding-window approach (20 s windows, 7 s shift) was applied to the resting-state time series, followed by Fisher z-transformed correlations and a stepwise functional connectivity analysis. The resulting local and distant rs-dFC maps represented the primary neuroimaging outcomes, indexing the temporal consistency of short- and long-range functional interactions. Further details on image acquisition, preprocessing, and analytic procedures are available elsewhere ^16^.

### 2.5. Alzheimer’s Disease-risk-related rs-dFC peak values

Peak coordinates were taken from brain regions that our previous work had identified as showing altered local or distant rs-dFC associated with AD risk markers, including APOE4 genotype, amyloid-β burden, and plasma NfL levels ^16^; see Supplementary Table 1 for the selected peak coordinates and direction of the original associations). Building on this previous work, we extracted the Fisher z-transformed rs-dFC values at each of these peak coordinates from the individual connectivity maps of all participants. When local peaks overlapped spatially but originated from different AD-risk-related markers, only the more significant coordinate was retained to avoid redundancy. These selected AD-risk-related rs-dFC peak coordinates were then used as regions of interest (ROIs) in subsequent association analyses with gait variability and physical fitness indicators.

### 2.6. Data and statistical Analyses

The characteristics of the study sample are presented for the whole sample and stratified by sex, with continuous variables summarized as mean and standard deviation (SD). Normality of the study variables was assessed using the Kolmogorov–Smirnov test, histograms, and Q–Q plots. Missing data were present for four participants, who had insufficient gait data caused by data collection errors. Due to the low proportion of missing data, no imputation was performed.

#### Correlation analyses

Before conducting the main analyses, two correlation matrices were generated to examine the relationships between covariates (i.e., age, sex, and education) and the main study variables. The first matrix included covariates and the AD–risk-related rs-dFC peaks and the second included gait variability indicators, physical fitness indicators, and covariates.

#### Targeted analyses (AD-risk-related rs-dFC regions)

We conducted targeted analyses to determine whether gait variability and physical fitness indicators were related to the targeted AD-risk-related rs-dFC regions. To this end, separate linear regression models were performed with gait variability and physical fitness indicators as predictors and AD-risk-related local and distant rs-dFC peak coordinate values as dependent variables. All models were adjusted for age, sex and education. Multiple comparisons were controlled using the false discovery rate (FDR) correction ^22^, applied separately to the set of three local rs-dFC peaks and the set of eight distant rs-dFC peaks. Statistical significance for FDR-adjusted results was set at p<0.05 (two-tailed).

#### Whole-brain analyses

We employed separate linear regression models to examine the associations of gait variability and physical fitness indicators with local and distant rs-dFC maps. All models were adjusted for age, sex, and education. These analyses were conducted using Statistical Parametric Mapping software (SPM12; Wellcome Department of Cognitive Neurology, MA). Statistical significance was set at an uncorrected voxel-level threshold of P ≤ 0.01 and a cluster-level FWE-corrected pFWE < 0.05, with corrections applied within a gray matter mask derived from SPM12 tissue probability maps. Then, for each brain region identified in the voxel-wise analysis as showing a significant association with the physical performance indicator of interest, we extracted the Fisher z-transformed rs-dFC temporal consistency value at the peak voxel using SPM12. These extracted peak values were subsequently used to calculate standardized β coefficients in RStudio for Mac (Version 2023.03.1; lm.beta package).

#### Sensitivity analyses

For sensitivity analyses, we first fitted models adjusted for age, sex, and education (Model 1), and then additionally adjusted these models for body mass index (BMI) (Model 2). For gait variability analyses, Model 3 further included cardiorespiratory fitness (CRF; 2-km walking test), given its strong correlation with gait variability indicators. Finally, Model 4 additionally included APOE status on top of Model 2, to account for genetic burden, as genetic risk is commonly considered an important potential confounder in analyses of AD-related biomarkers and brain outcomes.

## 3. Results

### 3.1. Descriptive Characteristics

The descriptive characteristics of the study sample are presented in Table 1. The sample of 86 participants (60.5% females) had a mean age of 71.63 ± 3.94 years. The average education length was 11.53 ± 4.98 years. Supplementary Table 1 provides the descriptive values of the local and distant AD-risk-related rs-dFC peak coordinates.

**Table 1.** Demographic details of the study sample (n=86).

|  | Overall | Females | Males |
|---|---|---|---|
| **Overall characteristics** | 86 | 52 | 34 |
| Age, yr | 71.63 ± 3.94 | 71.87 ± 4.13 | 71.26 ± 3.68 |
| Education length, yr | 11.53 ± 4.98 | 9.75 ± 4.08 | 14.26 ± 5.05 |
| Weight, kg | 73.53 ± 13.67 | 67.48 ± 11.31 | 82.79 ± 11.74 |
| Height, cm | 160.44 ± 9.30 | 154.41 ± 4.10 | 169.68 ± 7.16 |
| Body-mass index, kg/m2 | 28.49 ± 4.36 | 28.36 ± 5.00 | 28.70 ± 3.20 |
| **Cognitive Status** |  |  |  |
| Montreal Cognitive Assessment, raw score | 25.53 ± 2.13 | 25.04 ± 2.14 | 26.29 ± 1.92 |
| Mini-Mental State Examination, raw score | 28.97 ± 1.07 | 28.98 ± 1.13 | 28.94 ± 0.98 |
| Telephone Interview of Cognitive Status, raw score | 33.97 ± 2.80 | 33.54 ± 2.86 | 34.62 ± 2.62 |
| **Gait variability indicators** |  |  |  |
| Stride length CV, % | 2.55 ± 0.98 | 2.79 ± 1.15 | 2.21 ± 0.50 |
| Gait velocity CV, % | 2.98 ± 0.97 | 3.19 ± 1.06 | 2.69 ± 0.75 |
| Step time CV, % | 2.58 ± 0.84 | 2.86 ± 0.89 | 2.19 ± 0.58 |
| Step length CV, % | 3.66 ± 1.40 | 4.12 ± 1.58 | 3.01 ± 0.73 |
| **Physical fitness** |  |  |  |
| ***Muscular strength*** |  |  |  |
| Handgrip strength, kg | 28.20 ± 9.93 | 21.11 ± 3.43 | 39.04 ± 5.93 |
| Biceps curl, rep | 16.01 ± 3.85 | 15.21 ± 3.76 | 17.24 ± 3.72 |
| Squats, rep | 13.98 ± 2.99 | 13.48 ± 2.87 | 14.74 ± 3.04 |
| Elbow extension strength, Nm | 90.83 ± 34.91 | 73.02 ± 21.96 | 118.06 ± 33.62 |
| Knee extension strength, Nm | 97.06 ± 34.76 | 77.11 ± 19.94 | 127.57 ± 30.19 |
| ***Cardiorespiratory fitness*** |  |  |  |
| 2-km walking test, min<sup>†</sup> | 23.26 ± 3.78 | 24.42 ± 3.83 | 21.50 ± 2.96 |
| 6-minutes walking test, m | 478.14 ± 73.97 | 456.40 ± 67.68 | 511.38 ± 71.64 |

*Notes:* CV = coefficient of variation; SD = standard deviation; rep = repetition; Nm = Newton × meter. Values are expressed as mean ± SD unless otherwise indicated. Gait variability indicators are expressed as CV ± SD; lower values indicate better gait stability. <sup>†</sup>Lower value means better cardiorespiratory fitness.

### 3.2. Correlation matrices

Correlation matrices among physical performance indicators, AD-risk–related rs-dFC regions, and covariates are presented in Fig. 1 and Supplementary Fig. 1.

**Fig. 1.**
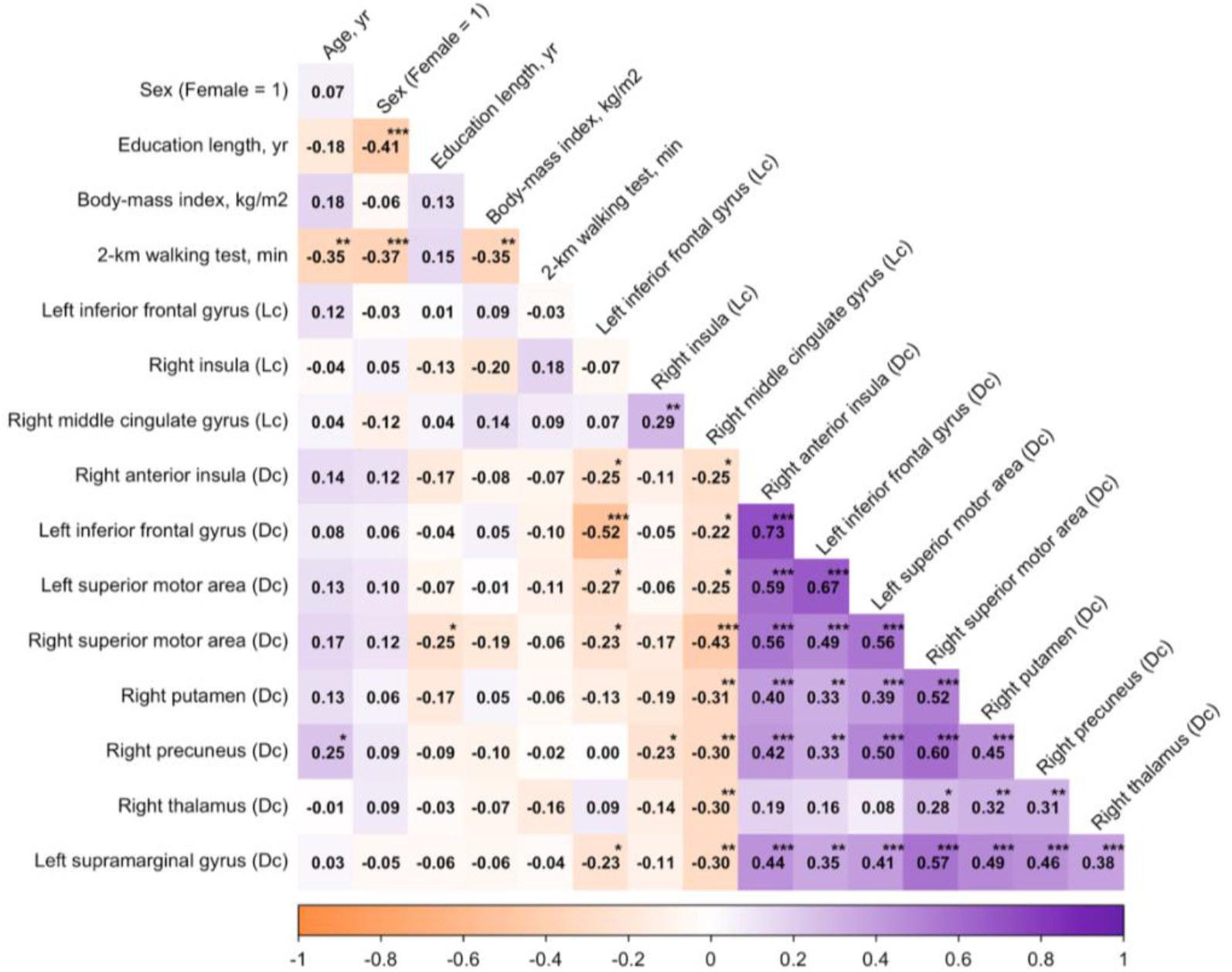
Correlations among AD-risk-related dFC peak values and covariates. Heatmap showing Pearson’s correlation coefficients (r) among covariates (age, sex, education, body-mass index and cardiorespiratory fitness) and AD-risk–related local and distant dFC peak values. Orange colors represent negative correlations, while purple colors indicate positive correlations. Values correspond to pairwise correlations, with darker shades reflecting stronger associations. Notes: LC = local connectivity; DC = distant connectivity. *p < 0.05, **p < 0.01, *** p < 0.001. Alt text: Triangular correlation heatmap showing relationships among demographic characteristics, body mass index, cardiorespiratory fitness, and Alzheimer’s disease risk-related local and distant dynamic functional connectivity values. Orange indicates negative correlations and purple indicates positive correlations, with darker shades representing stronger associations. Significant correlations are marked with asterisks. Several distant connectivity measures show positive correlations with one another.

Regarding rs-dFC measures, inter-regional correlations among local regions were generally weak and mostly non-significant (typically r < 0.30), with only isolated significant associations. In contrast, distant rs-dFC regions showed multiple significant positive intercorrelations (r ranging from 0.31 to 0.73). Correlations between local and distant rs-dFC measures were minimal (Fig. 1).

Gait variability and physical fitness indicators mostly showed significant inverse correlations (r ranging from −0.24 to −0.47). Covariates such as age, sex, education, and BMI were correlated with several gait and physical performance measures, with directions and magnitudes consistent across indicators (Supplementary Fig. 1).

### 3.3. Targeted analyses of association of physical performance indicators with AD-risk-related rs-dFC peak values

Fig. 2 and Supplementary Fig. 2 shows the associations between gait variability indicators and local and distant AD-risk-related rs-dFC peak values, controlling for age, sex and Education, respectively. Regarding local rs-dFC temporal consistency, greater stride length CV, gait velocity CV and step length CV were associated with greater local rs-dFC temporal consistency in the left inferior frontal gyrus (β ranging from 0.276 to 0.328, pFDR = 0.024 to 0.047). Conversely, greater gait velocity CV, step length CV and stride length CV were associated with lower local rs-dFC temporal consistency in the right insula (β ranging from - 0.473 to −0.313, pFDR = 0.001 to 0.028). No significant associations were found between step time CV and local rs-dFC temporal consistency (p > 0.05). Importantly, no significant associations emerged for distant rs-dFC temporal consistency across any gait variability parameter.

**Fig. 2.**
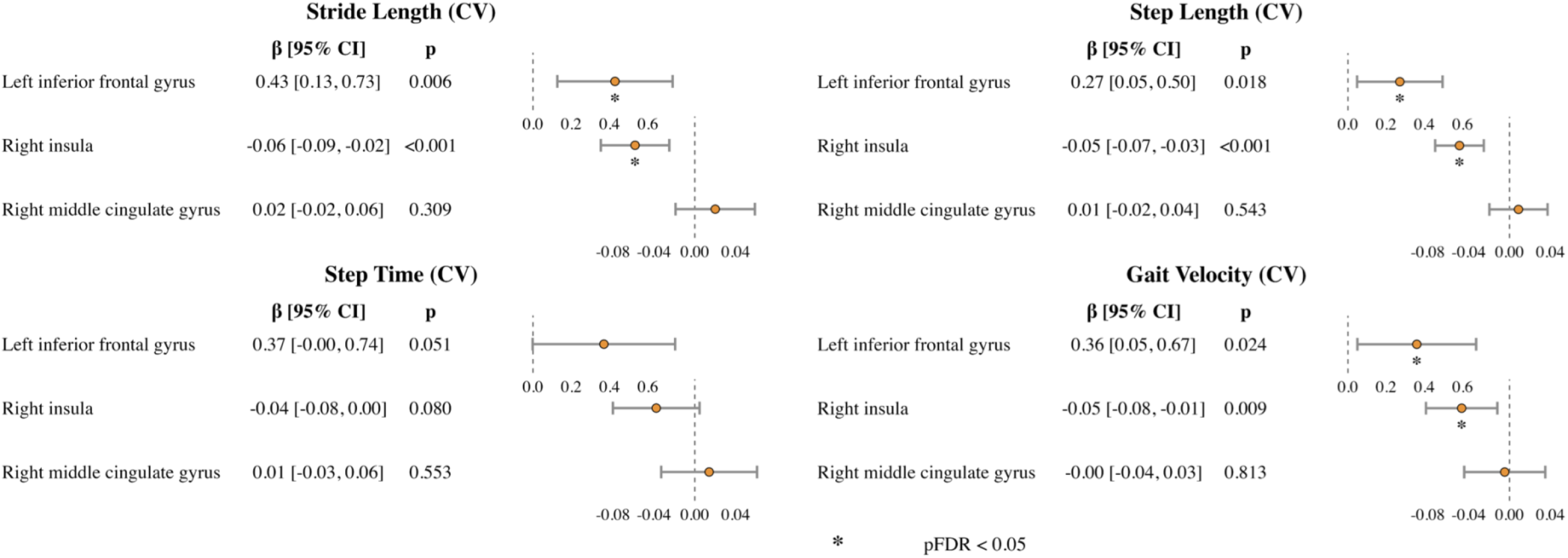
Associations between gait variability and Alzheimer’s disease risk–related local resting-state dynamic functional connectivity (rs-dFC) peak values. Forest plots display standardized regression coefficients (β) and 95% confidence intervals for the associations between gait variability measures and local rs-dFC peak values derived from predefined brain regions. All models were adjusted for age, sex, and years of education. Points represent standardized β estimates and horizontal lines indicate 95% confidence intervals. Asterisks indicate results surviving false discovery rate (FDR) correction. Two x-axis scales were used because confidence interval widths differed substantially across local peaks. Detailed information on the peak values, their associations with the biomarkers, and their interpretation can be found in Supplementary Table 1. Note. CV = coefficient of variation; rs-dFC = resting-state dynamic functional connectivity; β = standardized regression coefficient; CI = confidence interval; FDR = false discovery rate; p = p value. Local rs-dFC peak values represent Fisher z–transformed connectivity estimates. Alt text: Four forest plots showing associations between stride length, step length, step time, and gait velocity variability and local resting-state dynamic functional connectivity in three brain regions. Points represent standardized regression coefficients, and horizontal lines indicate 95% confidence intervals. Greater stride length, step length, and gait velocity variability are associated with higher local connectivity temporal consistency in the left inferior frontal gyrus and lower temporal consistency in the right insula. No significant associations are observed in the right middle cingulate gyrus. Asterisks identify associations surviving correction for multiple comparisons.

Supplementary Fig. 3 and 4 depict the non-significant associations observed between physical fitness indicators and AD-risk-related rs-dFC peak values after controlling for age, sex, and education (all pFDR > 0.05).

### 3.4. Whole-brain analyses of association of physical performance indicators with local and distant rs-dFC temporal consistency maps

Fig. 3 shows the associations between gait variability indicators and clusters located in brain regions with local and distant rs-dFC temporal consistency patterns, controlling for age, sex, and education. Regarding local rs-dFC temporal consistency, greater stride length CV was associated with lower local rs-dFC temporal consistency in the left parahippocampal gyrus (β = −0.428, k = 43, pFWE = 0.002). Greater gait velocity CV was also associated with lower local rs-dFC temporal consistency in the left calcarine cortex (β = −0.452, k = 46, pFWE = 0.001). No significant associations were found between step time CV or step length CV and local rs-dFC temporal consistency (p > 0.05). Importantly, no significant associations emerged for distant rs-dFC temporal consistency across any gait variability parameter.

**Fig. 3.**
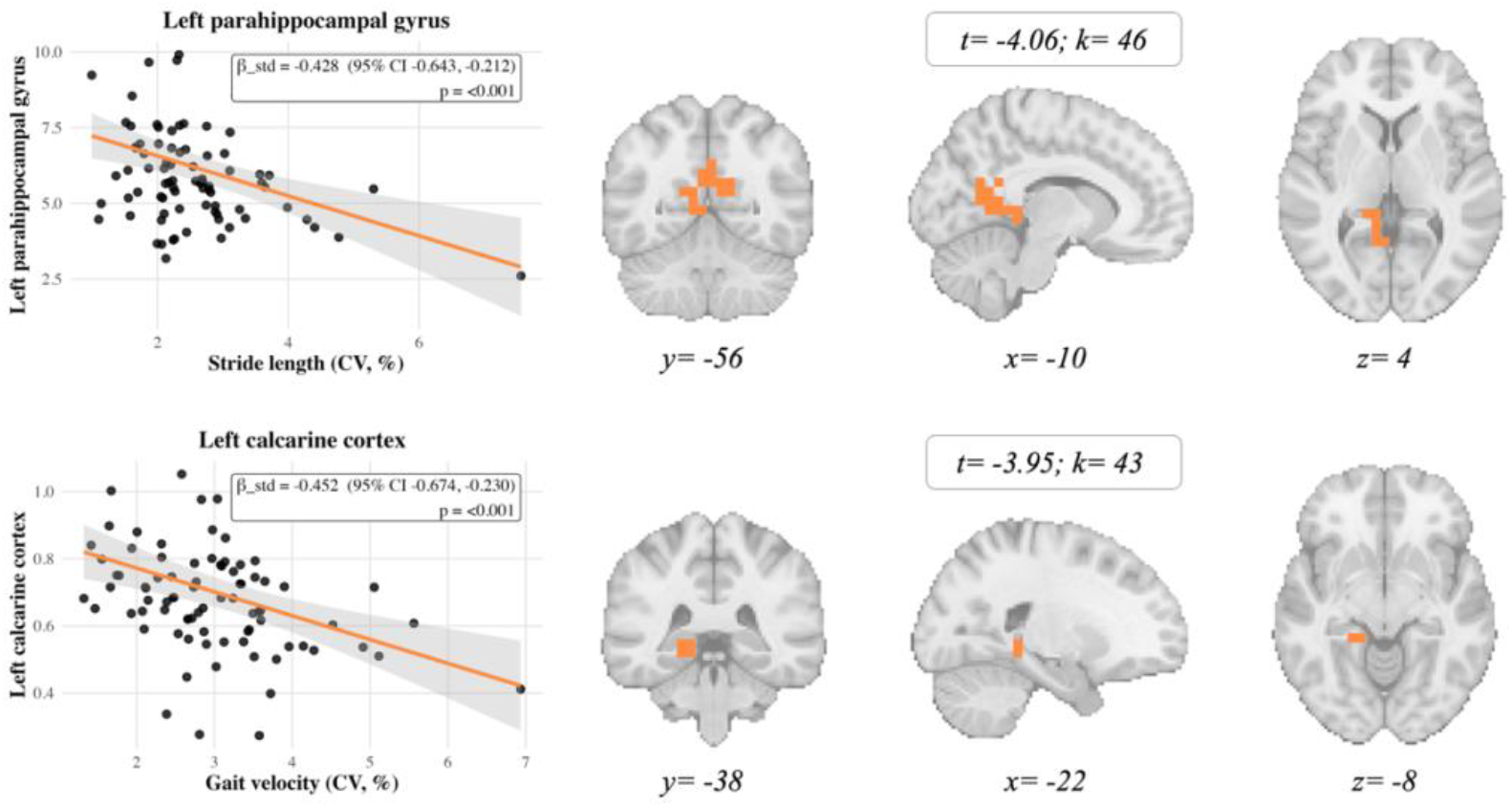
Associations between gait variability and local resting-state dynamic functional connectivity (rs-dFC). Scatter plots show the associations between gait variability measures and local rs-dFC peak values. No associations were revealed between gait variability and distant rs-dFC peak values. Solid lines represent adjusted linear regression fits, and shaded areas indicate 95% confidence intervals. All analyses were adjusted for age, sex, and years of education. Local rs-dFC peak values represent Fisher z–transformed connectivity estimates. Whole-brain statistical maps were thresholded at p < .01 (uncorrected) for display purposes. Brain images display the corresponding whole-brain clusters derived from voxel-wise analyses, with peak clusters highlighted in orange. Images are shown in neurological convention, with the left hemisphere displayed on the left and the right hemisphere on the right (L → R). *Note.* CI = confidence interval; CV = coefficient of variation; k = cluster size; t = strength and direction of the association; rs-dFC = resting-state dynamic functional connectivity; β = standardized regression coefficient. Alt text: Two sets of scatter plots and brain images showing negative associations between gait variability and local resting-state dynamic functional connectivity. The upper panel shows greater stride length variability associated with lower temporal consistency in the left parahippocampal gyrus. The lower panel shows greater gait velocity variability associated with lower temporal consistency in the left calcarine cortex. Scatter plots display individual observations, fitted regression lines, and 95% confidence intervals. Coronal, sagittal, and axial brain slices highlight the corresponding clusters in orange.

No significant associations were observed between any physical fitness indicators and either local or distant rs-dFC temporal consistency after controlling for age, sex, and education. (all pFWE > 0.05, all p-uncorrected > 0.01).

### 3.5. Sensitivity analyses controlling for BMI, CRF and APOE4

Supplementary Tables 2 reports the results of the sensitivity analyses. Overall, standardized regression coefficients remained highly similar across models, although some associations did not retain statistical significance after additional adjustment.

## 4. Discussion

The main finding of this study is that higher gait variability—but not other physical performance indicators such as muscular strength or CRF—was associated with rs-dFC temporal consistency in cognitively normal older adults. Importantly, these associations were observed specifically for local rs-dFC temporal consistency, whereas no associations were found for distant rs-dFC temporal consistency.

Local dynamic functional connectivity reflects the temporal consistency of communication among functionally close brain regions, indexing how consistently specialized neural systems coordinate their activity over time ^12–14^. Greater temporal consistency in local connectivity is often taken to reflect more reliable short-range coordination, whereas lower temporal consistency may indicate less consistent local coupling. Importantly, higher temporal consistency is not inherently beneficial or detrimental; rather, its functional meaning is context-dependent and may reflect either efficient, stable coordination or reduced flexibility depending on the system and behavioral demands ^23^. The selective association between gait variability—reflecting the consistency and regularity of walking patterns across steps—and the temporal consistency of local, but not distant, connectivity suggests that gait-related neural alterations in healthy aging primarily involve locally specialized network dynamics rather than long-range integration across distributed brain systems. This pattern is consistent with previous evidence showing that aging is characterized by reduced temporal consistency of local functional interactions, particularly within motor and cognitive control networks ^15^, and supports the idea that gait variability reflects early changes in the efficiency of local neural communication supporting locomotor control.

Within regions previously linked to AD risk markers, gait variability showed a consistent but region-specific pattern of associations with temporal consistency in local connectivity. Greater gait variability across stride length, gait velocity, and step length were associated with greater temporal consistency in local connectivity in the left inferior frontal gyrus, but lower temporal consistency in local connectivity in the right insula. The inferior frontal gyrus is a key region for executive control and top-down regulation of motor behavior ^24^. Greater temporal consistency of local connectivity in this region may therefore reflect a pattern consistent with increased involvement of executive control systems, in line with prior studies showing greater frontal recruitment during complex or unstable walking in older adults ^4,6,25^. In contrast, the insula plays a central role in multisensory and interoceptive integration, supporting the coordination of internal bodily signals with motor output ^26^. Lower temporal consistency of local connectivity in this region may indicate less stable sensorimotor integration, a process that has been shown to decline with aging and to relate to gait instability ^2,4^. Together, these complementary patterns suggest that gait variability may be associated with region-specific alterations in local functional organization involving executive-control and sensorimotor integration systems.

Whole brain analyses further supported this interpretation, showing that greater gait variability was associated with lower temporal consistency in local connectivity in regions involved in visuospatial processing and navigation, including the parahippocampal gyrus and the calcarine cortex. The parahippocampal gyrus is critically involved in spatial navigation and contextual processing ^27^, while the calcarine cortex contributes to visuomotor integration during locomotion ^28,29^. Reduced temporal consistency in local connectivity in these regions may reflect less temporally consistent coordination within cortical systems that integrate visual, spatial, and motor information, processes that are essential for adaptive gait control and are known to be vulnerable to aging ^6^. Importantly, these associations did not interact with AD–related genetic, pathological, or neurodegenerative markers previously examined in this cohor ^16^, suggesting that the observed rs-dFC temporal consistency patterns reflect age-related neural characteristics rather than AD–specific processes.

In contrast to gait variability, no associations were observed between muscular strength or CRF and temporal consistency in either local or distant connectivity. This finding suggests that different physical performance indicators reflect distinct aspects of brain–body coupling in aging. While gait stability appears to capture both functional and structural neural integrity ^19^, previous evidence suggests that CRF is more closely related to large-scale network organization and functional integration ^8,9,30^, and may be especially relevant for hub-based network properties that are metabolically costly to maintain ^31^. Similarly, associations between muscular strength and brain health have been more commonly described using static functional connectivity or structural brain measures ^11^. Supporting this distinction, prior work in the same cohort showed that poorer gait stability was associated with lower gray matter volume in hippocampal and parietal regions involved in spatial navigation and cognitive control ^19^, whereas muscular strength and CRF were associated with structural brain measures and executive function performance, often through inverse or region-specific associations rather than consistent positive effects ^20^. Together, these findings suggest that gait stability may be particularly sensitive to both dynamic functional processes and underlying brain structure ^19^, whereas muscular strength and CRF may relate more strongly to large-scale network organization or structural brain properties than to the temporal consistency of rs-dFC measured here ^20^.

Limitations should be acknowledged. First, the cross-sectional nature of the study precludes causal interpretations regarding the directionality of the observed associations; and second, the modest sample size may limit statistical power, particularly for detecting small effect sizes. Additionally, the inclusion of cognitively normal, physically inactive older adults aged 65–80 years may limit the generalisability of these findings to older populations with different cognitive or physical activity profiles. Despite these limitations, the study has several notable strengths. It applies an innovative analytic framework by combining gait variability and physical fitness indicators with dynamic functional connectivity—variables that have been scarcely examined in this context. Furthermore, it integrates both hypothesis-driven and whole-brain exploratory approaches and includes adjustment for key confounders, such as BMI, CRF and APOE4.

In summary, this study shows that greater gait variability, but not muscular strength or cardiorespiratory fitness, was associated with local rs-dFC temporal consistency in cognitively normal older adults. Specifically, gait variability related to temporal consistency in regions supporting executive control, sensorimotor integration, and visuospatial processing, suggesting that gait variability may be a sensitive marker of local neural coordination relevant to locomotor control. In contrast, the absence of associations for muscular strength and CRF suggests that these physical fitness indicators may be less tightly coupled to the temporal consistency of rs-dFC at rest, and may instead relate more strongly to structural brain properties or large-scale network organization than to the dynamic functional coordination indexed by rs-dFC. Future longitudinal and intervention studies with larger samples should test whether improvements in gait and other fitness domains lead to changes in rs-dFC dynamics over time, and whether distinct physical fitness indicators map onto complementary neural mechanisms supporting mobility and cognitive health in aging.

## Data Availability

The datasets generated and/or analyzed during the current study are not publicly available due to ethical and privacy considerations related to participant data but may be made available from the corresponding author upon reasonable request, subject to applicable ethical approvals and data-sharing agreements.

## Acknowledgements

The authors gratefully acknowledge all individuals who contributed to the conduct of this study and supported its completion. The authors also thank the participants of the AGUEDA trial for their time, commitment, and valuable contribution to this research.

## Declaration of generative AI use

During the preparation of this manuscript, the authors used ChatGPT (OpenAI) solely to assist with language editing and improve the clarity and readability of the text. All suggested changes were reviewed and approved by the authors, who take full responsibility for the final content of the manuscript.

## Supplementary Material

### Online Resource 1. Supplementary figures and tables

Supplementary material containing four figures illustrating correlations among gait variability, physical fitness, and demographic characteristics, as well as associations of gait variability and physical fitness with local and distant resting-state dynamic functional connectivity. Two supplementary tables provide information on Alzheimer’s disease risk-related connectivity peaks and additional regression models examining gait variability–brain associations.

**Supplementary Fig. 1.**
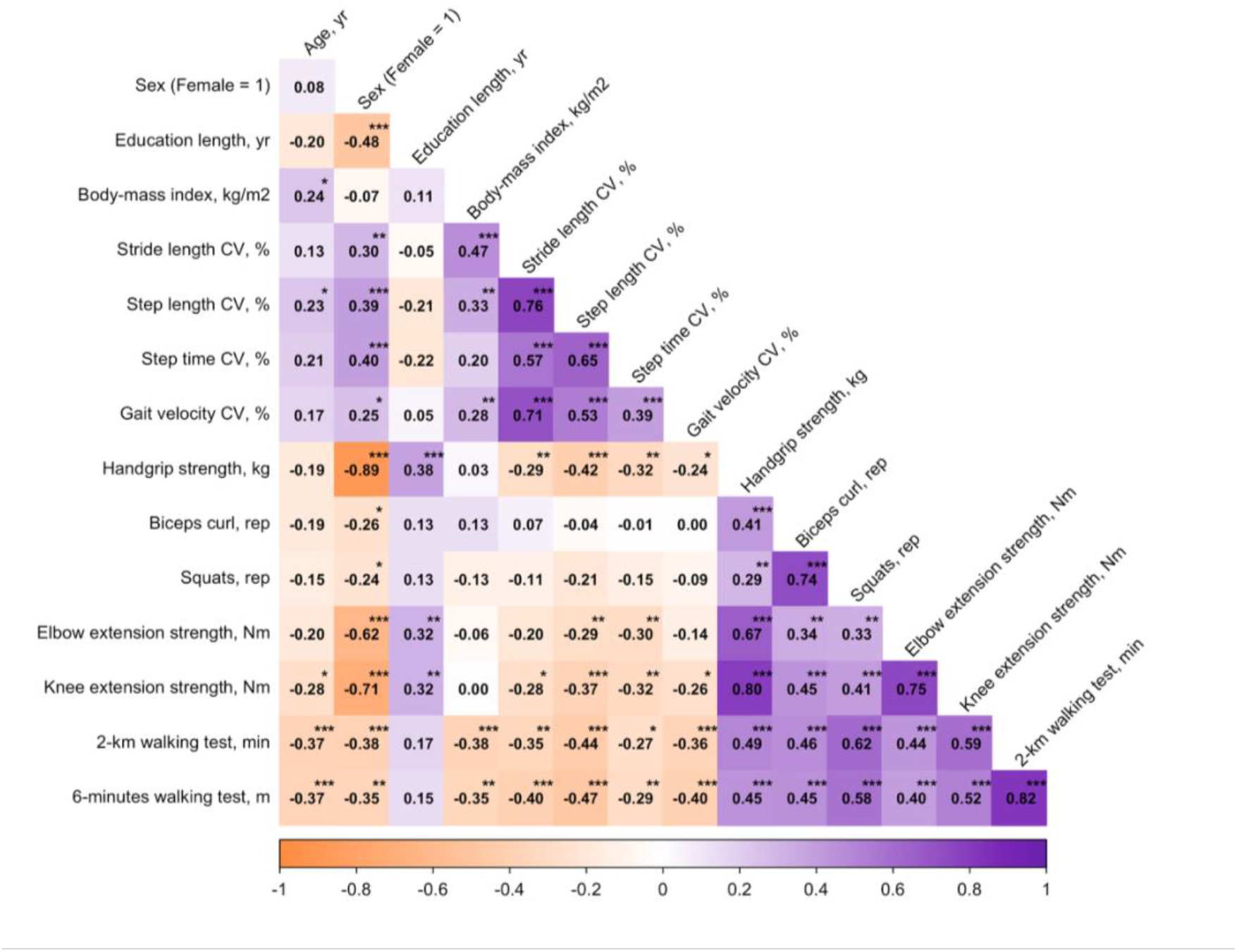
Correlations among gait variability indicators, physical fitness indicators, and covariates. Heatmap depicting Pearson’s pairwise correlation coefficients (r) among gait variability indicators (stride length, step length, step time, and gait velocity coefficients of variation), physical fitness indicators (handgrip strength, biceps curl, squats, elbow and knee extension strength, 2-km walking test, and 6-minute walking test), and demographic covariates (age, sex, education length, and body-mass index). Orange colors represent negative correlations, while purple colors indicate positive correlations, with darker shades reflecting stronger associations. Notes: CV = coefficient of variation; rep = repetitions; yr = years; kg/m² = kilograms per square meter; m = meters. *p < 0.05, **p < 0.01, ***p < 0.001.

**Supplementary Fig. 2.**
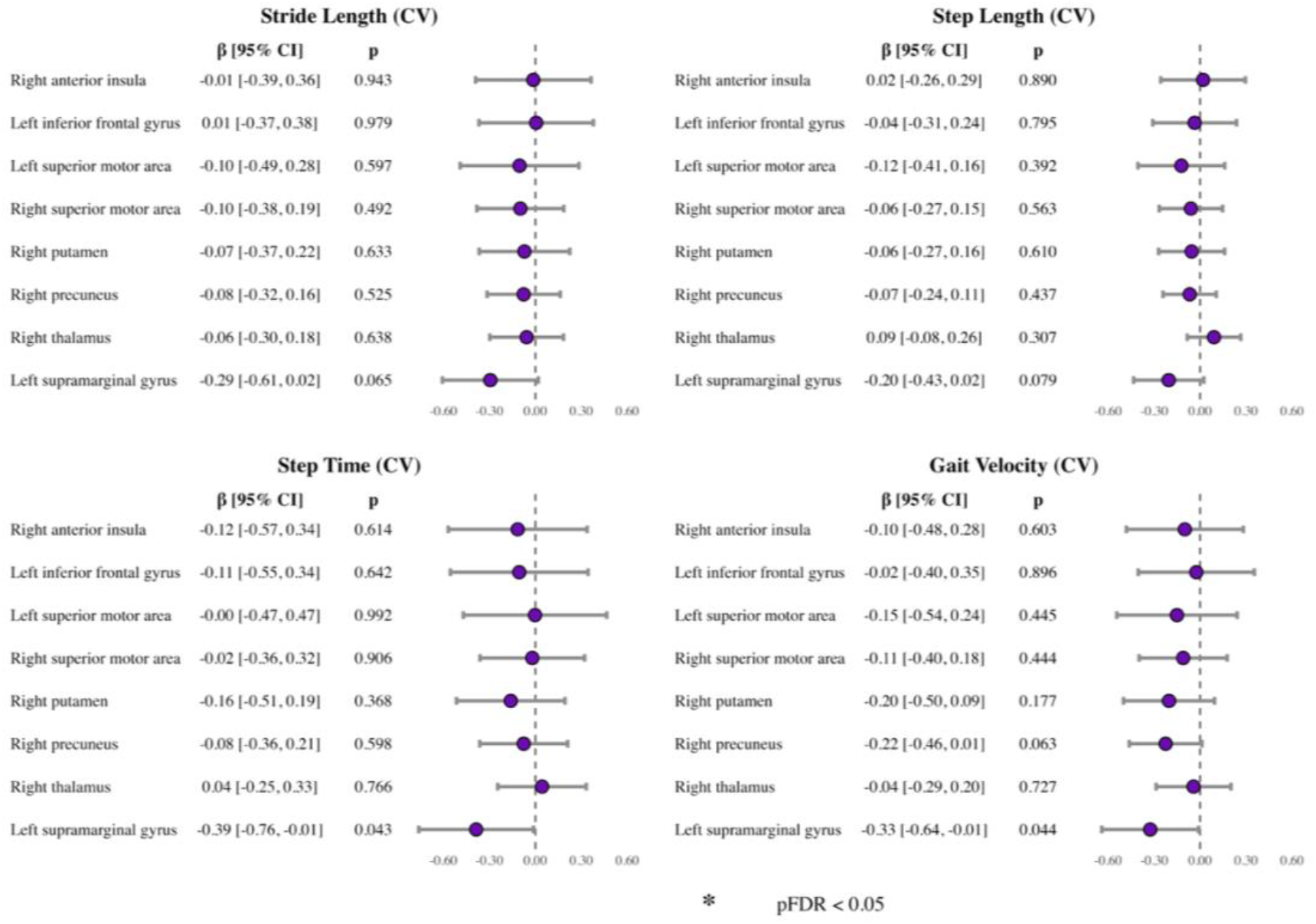
Associations between gait variability and Alzheimer’s disease risk–related distant resting-state dynamic functional connectivity (rs-dFC) peak values. Forest plots display standardized regression coefficients (β) and 95% confidence intervals for the associations between gait variability measures and distant rs-dFC peak values derived from predefined brain regions. All models were adjusted for age, sex, and years of education. Points represent standardized β estimates and horizontal lines indicate 95% confidence intervals. Asterisks indicate results surviving false discovery rate (FDR) correction. Detailed information on the peak values, their associations with the biomarkers, and their interpretation can be found in Supplementary Table 1. Note. CV = coefficient of variation; rs-dFC = resting-state dynamic functional connectivity; β = standardized regression coefficient; CI = confidence interval; FDR = false discovery rate; p = p value. Distant rs-dFC peak values represent Fisher z–transformed connectivity estimates.

**Supplementary Fig. 3.**
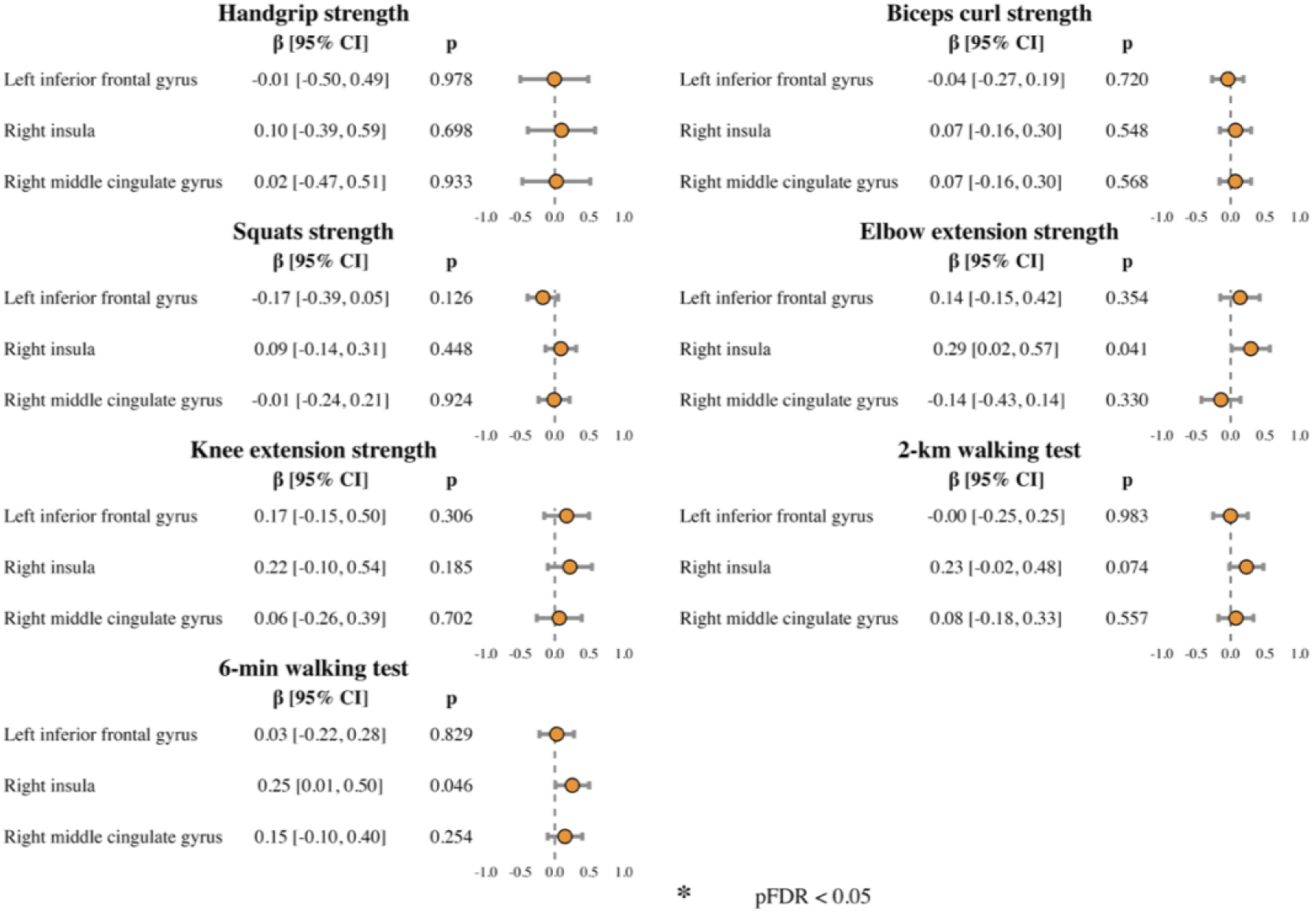
Associations between physical fitness and Alzheimer’s disease risk–related local resting-state dynamic functional connectivity (rs-dFC) peak values. Forest plots display standardized regression coefficients (β) and 95% confidence intervals for the associations between physical fitness measures and local rs-dFC peak values derived from predefined brain regions. All models were adjusted for age, sex, and years of education. Asterisks indicate results surviving false discovery rate (FDR) correction. Detailed information on the peak values, their associations with the biomarkers, and their interpretation can be found in Supplementary Table 1. Note. rs-dFC = resting-state dynamic functional connectivity; β = standardized regression coefficient; CI = confidence interval; FDR = false discovery rate. Local rs-dFC peak values represent Fisher z–transformed connectivity estimates.

**Supplementary Fig. 4.**
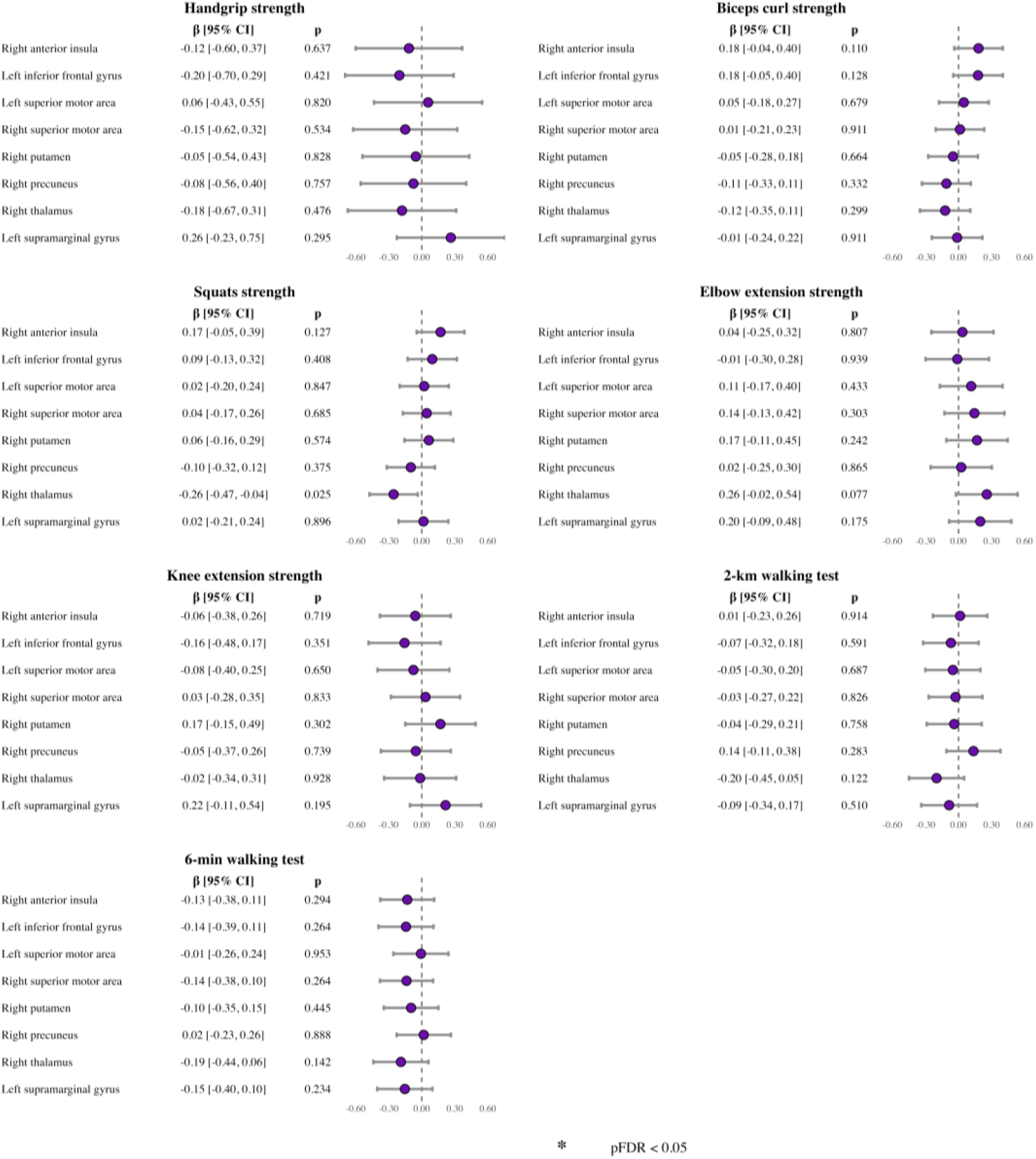
Associations between physical fitness and Alzheimer’s disease risk–related distant resting-state dynamic functional connectivity (rs-dFC) peak values. Forest plots display standardized regression coefficients (β) and 95% confidence intervals for the associations between physical fitness measures and distant rs-dFC peak values derived from predefined brain regions. All models were adjusted for age, sex, and years of education. Asterisks indicate results surviving false discovery rate (FDR) correction. Detailed information on the peak values, their associations with the biomarkers, and their interpretation can be found in Supplementary Table 1. Note. rs-dFC = resting-state dynamic functional connectivity; β = standardized regression coefficient; CI = confidence interval; FDR = false discovery rate. Distant rs-dFC peak values represent Fisher z–transformed connectivity estimates.

**Supplementary Table 1.** AD-risk-related rs-dFC peak values and coordinates selected for the targeted analyses.

| Brain Region | x | y | z | Connectivity Type | Direction Relative to AD Risk | Overall | Female | Male |
|---|---|---|---|---|---|---|---|---|
| Inferior frontal gyrus | -52 | 10 | 22 | Local | APOE4 carriers associated with lower local rs-dFC | 5.99 ± 1.34 | 5.95 ± 1.39 | 6.04 ± 1.27 |
| Inferior frontal gyrus | -4 | 4 | 58 | Distant | APOE4 carriers associated with higher distant rs-dFC | 4.78 ± 1.49 | 4.85 ± 1.50 | 4.67 ± 1.49 |
| Anterior Insula | -52 | 10 | 16 | Distant | APOE4 carriers associated with higher distant rs-dFC | 4.52 ± 1.52 | 4.68 ± 1.48 | 4.27 ± 1.57 |
| Superior Motor Area | -4 | 4 | 58 | Distant | APOE4 carriers associated with higher distant rs-dFC | 4.61 ± 1.61 | 4.73 ± 1.55 | 4.42 ± 1.69 |
| Middle Cingulate | 2 | -8 | 40 | Local | Higher plasma levels of NfL associated with lower local rs-dFC | 0.64 ± 0.16 | 0.63 ± 0.14 | 0.66 ± 0.18 |
| Insula | 38 | -14 | 16 | Local | Higher plasma levels of NfL associated with lower local rs-dFC | 0.65 ± 0.15 | 0.66 ± 0.14 | 0.64 ± 0.15 |
| Precuneus | 2 | -56 | 52 | Distant | Higher plasma levels of NfL associated with higher distant rs-dFC | 4.36 ± 1.00 | 4.43 ± 0.99 | 4.24 ± 1.01 |
| Putamen | 32 | -2 | 4 | Distant | Higher plasma levels of NfL associated with higher distant rs-dFC | 4.00 ± 1.20 | 4.06 ± 1.19 | 3.91 ± 1.23 |
| Superior Motor Area | 8 | -2 | 46 | Distant | Higher plasma levels of NfL associated with higher distant rs-dFC | 4.21 ± 1.20 | 4.34 ± 1.33 | 4.00 ± 0.95 |
| Supramarginal | -64 | -38 | 28 | Distant | Higher plasma levels of NfL associated with higher distant rs-dFC | 4.92 ± 1.31 | 4.86 ± 1.42 | 5.00 ± 1.13 |
| Thalamus | 2 | -8 | 10 | Distant | Higher plasma levels of NfL associated with higher distant rs-dFC | 4.76 ± 0.97 | 4.84 ± 0.94 | 4.64 ± 1.02 |

Note. Values are expressed as mean ± SD and represent Fisher’s z-transformed resting-state dynamic functional connectivity (rs-dFC) values extracted from AD-risk-related local and distant integration peaks. Higher Fisher’s z values indicate stronger functional connectivity. Peak coordinates are reported in MNI space and were selected from previous work showing significant associations between AD risk markers and rs-dFC in cognitively normal older adults. The column Direction Relative to AD Risk indicates whether higher AD risk was associated with lower or higher rs-dFC at each selected peak. When local peaks overlapped spatially across AD risk markers, only the more significant coordinate was retained to avoid redundancy. Sample size varied slightly across peak variables: APOE4-derived peaks were available for 84 participants (51 females, 33 males), whereas NfL-derived peaks were available for 85 participants (52 females, 33 males). Abbreviations: dFC, dynamic functional connectivity; SD, standard deviation; NfL, neurofilament light. Source: Coca-Pulido et al., 2026.

**Supplementary Table 2.** Associations between gait variability indicators and AD-risk-related local peak values derived from resting-state dynamic functional connectivity in cognitively normal older adults across four regression models.

| **Gait indicators** | **Brain region** | **β Model 1** | **β Model 2** | **β Model 3** | **β Model 4** |
|---|---|---|---|---|---|
| *Targeted analyses (AD-risk-related rs-dFC local peak values)* |  |  |  |  |  |
| Stride length CV | Left inferior frontal gyrus | **0.328** | **0.383** | **0.401** | **0.421** |
|  | Right insula | **-0.391** | **-0.450** | **-0.441** | **-0.506** |
| Gait velocity CV | Left inferior frontal gyrus | **0.276** | 0.268 | 0.295 | **0.294** |
|  | Right insula | **-0.313** | **-0.306** | -0.286 | **-0.326** |
| Step length CV | Left inferior frontal gyrus | **0.299** | 0.306 | 0.345 | **0.324** |
|  | Right insula | **-0.473** | **-0.499** | **-0.482** | **-0.551** |
| *Whole-brain analyses* |  |  |  |  |  |
| Stride length CV | Left parahippocampal gyrus | **-0.428** | -0.411 | -0.406 | **-0.063** |
| Gait velocity CV | Left calcarine cortex | **-0.452** | -0.398 | -0.394 | **-0.755** |

Linear regression models were conducted to examine the associations between gait variability indicators (predictor variables) and AD-risk–related local peak values derived from temporally consistent dynamic functional connectivity (dFC) patterns. Model 1 was adjusted for age, sex, and education; Model 2 additionally included BMI; Model 3 further included cardiorespiratory fitness (2-km walk test); and Model 4 additionally included APOE status on top of Model 2, to account for genetic burden as an important factor in AD-related analyses. Statistically significant associations after false discovery rate correction (pFDR < 0.05) are shown in bold. β = standardized regression coefficient. Lower values of gait variability indicators indicate better performance.
*Note.* AD-risk–related local peak values were extracted from individual local rs-dFC temporal consistency maps (Fisher z-transformed). pFDR indicates false discovery rate–adjusted p-values (Benjamini–Hochberg). CV = coefficient of variation; BMI = body mass index; CRF = cardiorespiratory fitness (2-km walk test, inverted so higher values indicate better fitness); APOE = apolipoprotein E genotype/status.

